# Non-pharmaceutical interventions and vaccinating school children required to contain SARS-CoV-2 Delta variant outbreaks in Australia: a modelling analysis

**DOI:** 10.1101/2021.10.03.21264492

**Authors:** George J Milne, Julian Carrivick, David Whyatt

## Abstract

**Background:** In countries with high COVID-19 vaccination rates the SARS-CoV-2 Delta variant has resulted in rapidly increasing case numbers. This study evaluated the use of non-pharmaceutical interventions (NPIs) coupled with alternative COVID-19 vaccination strategies to determine feasible Delta mitigation strategies for Australia. We aimed to understanding the interplay between high vaccine coverage levels and NPI physical distancing activation, and to establish the benefit of adding children and adolescents to the vaccination program. Border closure has limited SARS-CoV-2 transmission in Australia, however slow vaccination uptake resulted in Delta outbreaks in the two largest cities, and may continue once international travel resumes.

**Methods:** An agent-based model was used to evaluate the potential reduction in the COVID-19 health burden resulting from alternative vaccination strategies. We assumed immunity was derived from vaccination with the BNT162b2 Pfizer BioNTech vaccine. Two age-specific vaccination strategies were evaluated, age 5 and above, and 12 and above, and the health burden determined under alternative coverages, with/without activation of NPIs. Age-specific infections generated by the model, together with recent UK data, permitted reductions in the health burden to be quantified.

**Results:** Cases, hospitalisations and deaths are shown to reduce by: *i*) increasing coverage to include children aged 5 to 11 years, *ii*) activating moderate NPI measures, and/or *iii*) increasing coverage levels above 80%. At 80% coverage, vaccinating ages 12 plus without NPIs is predicted to give 1,162 hospitalisations; adding ages 5 and above gives 1,073 cases per million population. Activating moderate NPIs reduces hospitalisations to 705. Alternatively, increasing coverage to 90% reduces this to 684. Combining all three measures is shown to reduce cases to 398, hospitalisations to 2 and deaths to zero.

**Conclusions:** Delta variant outbreaks may be successfully managed by an achievable 80% vaccine coverage rate and moderate NPI measures, allowing schools and many workplaces to remain open. This prevents use of hard lockdown measures, and consequential economic and societal damage. Activating moderate NPIs is shown to give a similar reduction in health burden as increasing coverage of ages 12 and above to 90%. If 90% coverage cannot be achieved, including children and adolescents in the vaccination program coupled with moderate NPIs appears necessary to contain future COVID-19 Delta transmission.

**Funding:** The authors acknowledge funding support from the Department of Health, Western Australia (Future Health Research and Innovation Fund) and the Department of Health, Queensland, Australia.

## Background

By May 2021 countries which had vaccinated significant proportions of their population, such as Israel, the UK and the USA, saw significant reductions in daily diagnosed SARS-CoV-2 case numbers.^1^ From June 2021 onwards the more transmissible B1.617.2 Delta variant became dominant worldwide, even in countries with high vaccination rates.^2^ Given the rapid rise in Delta case numbers, the addition of children to COVID-19 vaccination schedules is under discussion by health authorities. Our model-based study aimed at understanding how expanding vaccination to younger age groups may reduce and contain this increase in case numbers, and consequential pressure on health systems.

Mathematical modelling has been used successfully to inform vaccination policy, providing evidence on effectiveness of alternative vaccination strategies, such as the benefit of increasing influenza vaccination in children.^3,4^ We evaluated the similar role childhood vaccination may have in a COVID-19 Delta context, and determined vaccine coverage levels needed to reduce and prevent growth in case numbers, with and without reintroduction of strict social distancing measures, as has occurred in Australia in 2020 and 2021.^5^

## Methods

An individual-based model capturing the demographics and movement patterns of individuals within the Australian city of Newcastle (population ∼273,000), together with SARS-CoV-2 virus transmission data from the early outbreak in Wuhan, China prior to social distancing activation ^6^, was previously developed. This model was used to analyse effectiveness of non-pharmaceutical social distancing interventions, varying their stringency, timing, and duration.^7,8^ In this study the probability of transmission occurring between infectious/susceptible pairs of individuals was adjusted to give a basic reproduction R_0_ = 6.0 for the COVID-19 Delta variant (95% CI [5.8, 6.1]).^2^

Census data captured age-specific demographics of every individual in each household in the modelled community, assigning adults to workplaces, and children to age-specific classes.^9-12^ The agent-based modelling technology captures the daily time-changing contact patterns of individuals, and the physical virus transmission “world” simulated by running model software using methods applied previously.^3,4,8^ Infection data generated by the model was translated into cases, hospitalisations and deaths using recent UK Delta variant data^13,14^ and a per million value was calculated to permit results to be scaled to population centres of different sizes. We modelled the BNT162b2 Pfizer vaccine with assumed protection against Delta infection of 88%. This percentage of individuals is assumed to remain immune for a period of at least 6 months, without immunity waning.^15^ Further model parameters are presented in Table 1.

**Table 1.** UWA COVID-19 model parameter settings

|  |  |
| --- | --- |
| **Model population** | ~273,000. 2011 census data for Newcastle and Lake Macquarie East, NSW. |
| **Assumed delta variant basic reproductive number** | 6.0 (95% CI [5.8, 6.1]) |
| **Time from infection to symptoms** | 5 days |
| **Time from infection to infectious** | 3 days |
| **Time from infection to recovery** | 9 days (4 days of symptoms, 6 days being infectious) |
| **Child (<12 years) susceptibility** | Same as adults |
| **Child (<12 years) transmissibility** | 50% of adults |
| **Adolescent (12-17 years) transmissibility** | Same as adults |
| **Probability of asymptomatic infections** | 35% |
| **Probability of asymptomatic transmission** | 55% of symptomatic transmission |
| **Probability of symptomatic infection diagnosis** | 33% |
| **Home isolation of diagnosed cases** | 7 days |
| **Seeded infections** | On average 1 infection every 4 days for 30 days |
| **BNT162b2 Pfizer vaccine** | Efficacy: 88% (against infection and transmission) |
| **Trigger for NPIs** | 1 diagnosed case in a single day |
| **Moderate NPIs** | **School closures:** 0%<br>**Workplace non-attendance:** 20%<br>**Community contact reduction:** 60% |
| **Strict NPIs** | **School closures:** 100%<br>**Workplace non-attendance:** 50%<br>**Community contact reduction:** 80% |

The short-term seeding of infectious individuals into the model was used to initiate a COVID-19 outbreak, at the rate of a single infection every 4 days for 30 days, equivalent to 4 infections seeded in a population of 1 million. The model captures the age of each modelled individual in one of 10 age bands, allowing age-specific health outcomes to be determined following infection.^16^ Model outputs captured the infection history of all individuals in the community, representing where and when infections occur. These analyses quantified infection and case reductions arising from increased vaccination coverage, and the reduction in hospitalisations and deaths.

We assumed use of the BNT162b2 (Pfizer) vaccine which affords protection against symptomatic disease by the Delta variant lower than that for the Wuhan strain,^7^ and the B.1.1.7 (Alpha) variant, reducing from 93% (Alpha) to 88% (Delta).^15^ In the absence of further evidence, we assumed protection against infection achieved to be the same as protection against disease. In the model the effect of vaccination is captured at the individual level, where a certain percentage of fully vaccinated individuals “move” from a susceptible state to an immune state four weeks post second-dose vaccination. The percentage of individuals moving to an immune state thus reflects vaccine efficacy.

The effectiveness of a particular vaccination strategy was determined by running model software after adjusting vaccination settings for alternative coverage levels and ages vaccinated. Seeding infectious individuals into the model initiated a COVID-19 Delta outbreak in a COVID-19 naive population.^8^ The median value of 100 simulation runs is presented; to aid clarity, confidence intervals are not presented, but were narrow. We assumed two doses of BNT162b2 Pfizer vaccine gave protection against Delta infection of 88%. Protection against poor health outcomes in those vaccinated and having breakthrough infections, was implicitly included by using recent UK Delta variant data^13^ to give age-specific case/hospitalisation and case/mortality ratios.

## Results

Results are presented for vaccination scenarios under alternative vaccine coverages, varied from 70% to 90% in individuals in two age categories, 5 and over and 12 and over. We also consider activating *moderate* physical distancing non-pharmaceutical (NPI) measures when Delta cases first appear in a community. These correspond to the Stage 3 measures adopted by the State of Victoria, Australia in 2020 and 2021, estimated as 30% workplace non-attendance, 75% community contact reduction, 70% case isolation^8^, altered to include schools remaining open, termed moderate NPIs in Table 1. Strict NPIs in Table 1 correspond to Victoria’s Stage 4 lockdown measures.

Table 2 presents an overview of results, allowing alternative vaccination strategies to be compared. These data suggest that increasing child-hood vaccinations to include ages 5 to 11, in addition to adolescents aged 12 and above, is significant. The rapid activation of strict physical distancing NPIs involving substantial economic, educational and societal disruption with schools closed (defined in Table 1) is shown in Table 2 to be highly effective in containing COVID-19 variants such as Delta, R_0_=6.0.

**Table 2.**
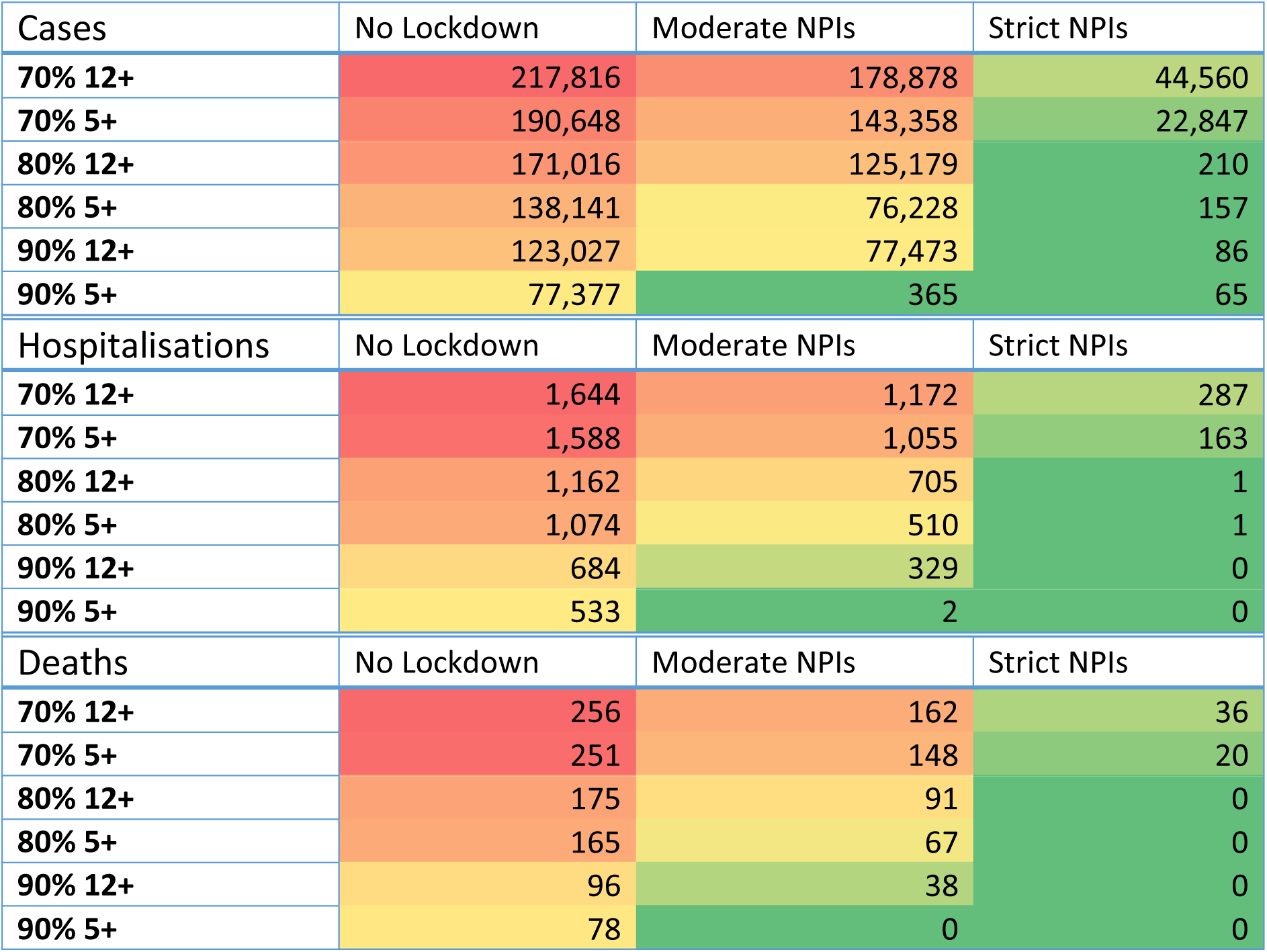
Total cases, hospitalisations and deaths per 1,000,000 population. Transmission calibrated to R_0_ of 6.0. Vaccine efficacy of 88% assumed for all ages. Median value of 100 simulations presented. Colours are linearly distributed according to the value along a minimum (green)/median (yellow)/maximum (red) scale.

In the absence of any physical distancing measures (moderate or strict NPIs) this strategy is shown in Figure 1 to reduce the peak in daily case numbers by approximately 2000 per 1 million population, for coverage levels of 70% and 80% of these alternative vaccination age groups. At 90% coverage, the benefit of including younger children is more pronounced, reducing the peak in cases by over half, from ∼3800 to ∼1500, as illustrated in the predicted epidemic curves, Figure 1.

**Figure 1.**
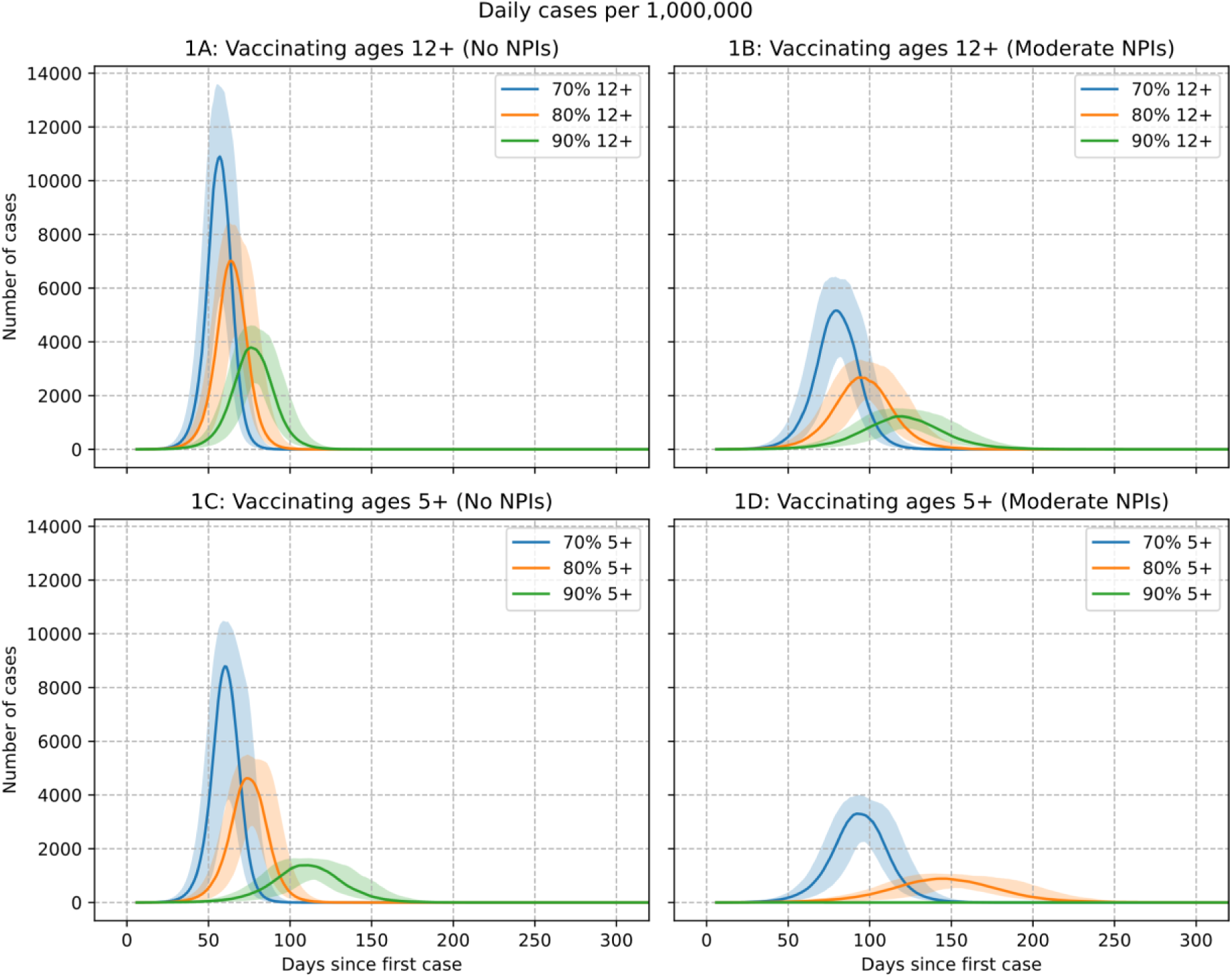
Daily cases per million for alternative vaccine coverage levels, with and without moderate NPIs. Vaccine with 88% efficacy assumed for all ages. R_0_ of=6.0. Median value of 100 simulations presented, with 10^th^ and 90^th^ percentile as shaded areas.

With the addition of moderate physical distancing measures, which include schools remaining open, data in Table 2 further highlights the benefit of vaccinating these younger age groups. At an achievable coverage level of 80%, the total number of cases in a population of 1 million, is reduced from ∼171,000 to ∼138,000 without any lockdown, and from 125,000 to 75,000 with moderate lockdown activated. The data illustrates the reduction in the COVID-19 health burden, and associated reduction in pressure on health systems, from vaccinating children aged 5 and over in terms of the reduction in hospitalisations and deaths. With 80% coverage, hospitalisations and deaths are predicted to reduce slightly without moderate physical distancing measures. With moderate measures added, hospitalisations are seen to reduce from 705 to 510 per million, while deaths are predicted to reduce from 91 to 67 per million.

With the addition of moderate physical distancing measures, which include schools remaining open, data in Table 2 further highlights the benefit of vaccinating these younger age groups. At an achievable coverage level of 80%, the total number of cases in a population of 1 million, is reduced from ∼171,000 to ∼138,000 without any lockdown, and from 125,000 to 75,000 with moderate lockdown activated. The data illustrates the reduction in the COVID-19 health burden, and associated reduction in pressure on health systems, from vaccinating children aged 5 and over in terms of the reduction in hospitalisations and deaths. With 80% coverage, hospitalisations and deaths are predicted to reduce slightly without moderate physical distancing measures. With moderate measures added, hospitalisations are seen to reduce from 705 to 510 per million, while deaths are predicted to reduce from 91 to 67 per million.

Predicted case data illustrated by the epidemic curves in Figure 1 indicate that altering vaccination policy to include ages 5 to 11, in addition to adolescents aged 12 and above, is also significant in terms of the daily pressure on healthcare resources. In the absence of physical distancing measures (Fig. 1A and 1C) this strategy reduces the peak in daily case numbers by approximately 2000 per 1 million population, for coverage levels of 70% and 80%. At an 80% coverage rate, a peak of ∼2300 cases with vaccination of years 12 and above is reduced to ∼900 maximum per day, if also vaccinating ages 5 and above. At 90% coverage, the benefit of including younger children is more pronounced, reducing the peak in cases by over half, from ∼3800 to ∼1500. With the addition of moderate physical distancing measures which include schools remaining open, Figure 1B and 1D highlight the benefit of vaccinating these younger age groups.

Tables 3, 4 and 5 present the reduction in the COVID-19 health burden by age group bands. Cases, hospitalisations and deaths are all reduced by: *i*) increasing vaccine coverage to include children aged 5 to 11 years, *ii*) activating moderate physical distancing measures, and/or *iii*) increasing coverage levels above 80%. To reduce pressure on health systems, hospitalisation rates may be reduced by combing all or some of these 3 strategies.

**Table 3.** Cases per million for increasing vaccination coverage levels (ages 12 and up, and 5 and up), with and without moderate NPIs. Vaccine with 88% efficacy assumed for all ages. R_0_ of=6.0. Median value of 100 simulations presented.

| Cases |  | Total | 0-12y | 13-24y | 25-44y | 45-64y | 65-79y | 80y+ |
| --- | --- | --- | --- | --- | --- | --- | --- | --- |
| No NPIs | 70% 12+ | 217,816 | 68,399 | 30,447 | 47,533 | 45,071 | 18,350 | 7,995 |
|  | 70% 5+ | 190,010 | 43,146 | 30,173 | 46,674 | 44,378 | 17,915 | 7,789 |
|  | 80% 12+ | 171,016 | 64,809 | 21,916 | 34,506 | 31,976 | 12,427 | 5,409 |
|  | 80% 5+ | 137,612 | 36,164 | 21,367 | 32,765 | 30,593 | 11,704 | 5,054 |
|  | 90% 12+ | 123,027 | 60,900 | 12,914 | 21,034 | 18,539 | 6,795 | 2,914 |
|  | 90% 5+ | 76,333 | 25,074 | 11,069 | 17,150 | 15,321 | 5,424 | 2,298 |
| Moderate NPIs | 70% 12+ | 178,878 | 61,533 | 25,864 | 39,993 | 35,484 | 11,310 | 4,631 |
|  | 70% 5+ | 142,872 | 3,387 | 24,713 | 36,630 | 33,079 | 10,367 | 4,194 |
|  | 80% 12+ | 125,179 | 54,149 | 15,837 | 25,234 | 21,214 | 6,249 | 2,533 |
|  | 80% 5+ | 74,983 | 2,175 | 12,788 | 18,599 | 16,165 | 4,562 | 1,829 |
|  | 90% 12+ | 77,473 | 1,939 | 6,801 | 12,180 | 9,153 | 2,511 | 1,026 |
|  | 90% 5+ | 398 | 13 | 53 | 84 | 73 | 17 | 6 |

**Table 4.** Hospitalisations per million for increasing vaccination coverage levels (ages 12 and up, and 5 and up), with and without moderate NPIs. Vaccine with 88% efficacy assumed for all ages. R_0_ of=6.0. Median value of 100 simulations presented.

| Hospitalisations |  | Total | 0-12y | 13-24y | 25-44y | 45-64y | 65-79y | 80y+ |
| --- | --- | --- | --- | --- | --- | --- | --- | --- |
| No NPIs | 70% 12+ | 1,644 | 66 | 47 | 305 | 437 | 373 | 415 |
|  | 70% 5+ | 1,584 | 39 | 47 | 299 | 431 | 364 | 404 |
|  | 80% 12+ | 1,162 | 63 | 34 | 221 | 310 | 252 | 281 |
|  | 80% 5+ | 1,073 | 32 | 33 | 210 | 297 | 238 | 262 |
|  | 90% 12+ | 684 | 60 | 20 | 135 | 180 | 138 | 151 |
|  | 90% 5+ | 528 | 23 | 17 | 110 | 149 | 110 | 119 |
| Moderate NPIs | 70% 12+ | 1,172 | 61 | 40 | 257 | 344 | 230 | 240 |
|  | 70% 5+ | 1,054 | 32 | 38 | 235 | 321 | 211 | 218 |
|  | 80% 12+ | 705 | 54 | 25 | 162 | 206 | 127 | 132 |
|  | 80% 5+ | 504 | 20 | 20 | 119 | 157 | 93 | 95 |
|  | 90% 12+ | 329 | 47 | 11 | 78 | 89 | 51 | 53 |
|  | 90% 5+ | 2 | 0 | 0 | 1 | 1 | 0 | 0 |

**Table 5.** Deaths per million for increasing vaccination coverage levels (ages 12 and up, and 5 and up), with and without moderate NPIs. Vaccine with 88% efficacy assumed for all ages. R_0_ of=6.0. Median value of 100 simulations presented.

| Deaths |  | Total | 0-12y | 13-24y | 25-44y | 45-64y | 65-79y | 80y+ |
| --- | --- | --- | --- | --- | --- | --- | --- | --- |
| No NPIs | 70% 12+ | 256 | 0 | 1 | 5 | 44 | 80 | 127 |
|  | 70% 5+ | 250 | 0 | 1 | 4 | 43 | 78 | 124 |
|  | 80% 12+ | 175 | 0 | 1 | 3 | 31 | 54 | 86 |
|  | 80% 5+ | 165 | 0 | 1 | 3 | 30 | 51 | 80 |
|  | 90% 12+ | 96 | 0 | 0 | 2 | 18 | 30 | 46 |
|  | 90% 5+ | 77 | 0 | 0 | 2 | 15 | 24 | 37 |
| Moderate NPIs | 70% 12+ | 162 | 0 | 1 | 4 | 34 | 49 | 74 |
|  | 70% 5+ | 148 | 0 | 1 | 4 | 32 | 45 | 67 |
|  | 80% 12+ | 91 | 0 | 1 | 2 | 20 | 27 | 40 |
|  | 80% 5+ | 67 | 0 | 0 | 2 | 16 | 20 | 29 |
|  | 90% 12+ | 38 | 0 | 0 | 1 | 9 | 11 | 16 |
|  | 90% 5+ | 0 | 0 | 0 | 0 | 0 | 0 | 0 |

At 80% coverage, vaccinating ages 12 plus without NPIs is predicted to result in 1,162 hospitalisations, adding ages 5 plus reduces this to 1,073, see Table 4. If moderate NPIs are applied at 80% coverage of ages 5 and above: case numbers half to 76,228; hospitalisations half to 510; and deaths reduce from 165 to 67, per million population.

From Table 2, at 80% coverage of ages 12 and above without moderate NPIs, two strategies may be adopted to reduce the health burden further; activating moderate NPIs, or increasing vaccine coverage. Activating moderate physical distancing measures is shown to reduce hospitalisations to 705. Alternatively, increasing coverage to 90% results in a reduction to 684 hospitalisations. Thus, activating moderate NPIs is seen to give a similar reduction in hospitalisations as increasing coverage of ages 12 and above to 90%.

Combining all three measures; *viz*. vaccination children aged 5 to 11, activating moderate NPIs, and 90% coverage is shown in Tables 2,3,4 and 5 to reduce cases to 365, hospitalisations to 2 and deaths to zero, per million of the population.

A key aim of COVID-19 mitigation is to both lessen the mortality rate, and the demand on the health system. With hospital bed resources occupied by COVID-19 patients there are significant effects more generally, negatively impacting access to healthcare for non COVID-19 conditions. Excess hospitalisation demand, illustrated in Figure 2, was estimated using a rolling 2-week sum of daily hospitalisations with average duration taken from recent UK data.^17^ Peak ICU bed demand can thus be determined using hospitalisation/ICU ratios. From Figure 2, and in the absence of further interventions, 80% vaccination coverage of ages 12 and above may result in peak hospital bed demand of ∼600 beds per million. However, adding moderate NPIs can reduce this demand to ∼200 beds per million. With the addition of ages 5-11 to the vaccination program, and 80% coverage, we estimate ∼500 beds per million without NPIs, reducing to ∼100 beds per million with moderate NPI measures added.

**Figure 2.**
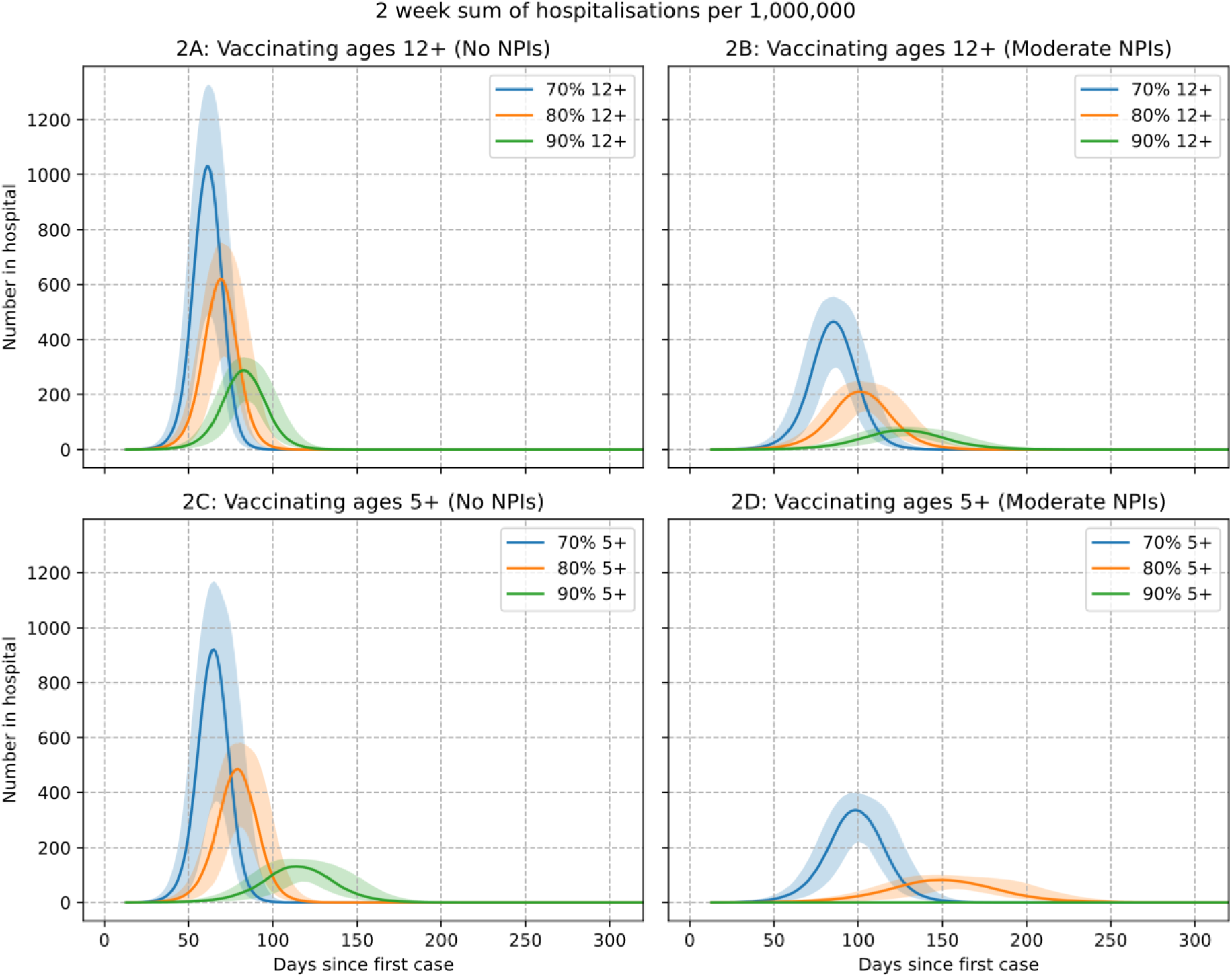
Two-week sum of hospitalisations per million population illustrating peak hospital demand for alternative vaccine coverage levels, with and without moderate NPIs. Median value of 100 simulations presented with 10^th^ and 90^th^ percentile shaded.

## Discussion

In the context of a highly transmissible SARS-CoV-2 variant, this study conducted a comprehensive modelling analysis of the population-wide effectiveness of a range of alternative vaccination strategies. These involved vaccination-only strategies, and those with attendant physical distancing measures activated, under a range of vaccine coverage levels. The aim was to determine vaccination strategies which best contain the spread of a highly transmissible SARS-CoV-2 variant (*i*.*e*. B1.617.2 Delta), reduce the resulting COVID-19 health burden, and lessen pressure on healthcare systems.

Key findings suggest that significant outbreaks of the SARS-CoV-2 Delta variant may continue to occur unless vaccination reaches high coverage levels in adults, adolescents and school-age children. Our study further suggests activation of physical distancing NPI measures will still be required, even in populations with high vaccination levels, and activating such measures as soon as case numbers start to increase. The addition of children aged 5 to 11 to the vaccination program was also found to be effective. Hard lockdown measures are known to cause societal and economic disruption, and significantly impact children’s education. However, moderate physical distancing measures with schools remaining open should be accepted by the public, and acceptable by health authorities given the reduction in the COVID-19 health burden resulting from such a policy. Results provide evidence that moderate lockdown measures with schools remaining open may successfully contain future high transmission variants under achievable coverage levels. They further highlight the need to activate such measures as early as possible. We determined that vaccinating adolescents and younger children was critical, to both increase the pool of immune individuals, and to mitigate transmission in an age group implicated in high transmission of viruses by aerosol droplets, as with influenza.^4^

The study assumed an Australian setting with effectively no naturally occurring immunity, given early border closures and adoption of a SARS-CoV-2 elimination policy. Herd immunity effects attained are the result of vaccination, rather than immunity derived from infection. Australia adopted an age-specific vaccination strategy in March 2021 due to limited supplies of mRNA vaccines, with those aged 60 year and over vaccinated with the AstraZeneca vaccine, manufactured locally. Our study assumed a modified vaccination strategy, using the Pfizer BioNTech mRNA vaccine to boost the immune response of those previously vaccinated with the AstraZeneca vaccine, necessary due to its lower efficacy against the Delta variant.^15,18-20^

The study quantified COVID-19 vaccination targets that may substantially reduce the growth in case numbers, and which avoid the need for economically and socially damaging hard lockdown measures. Adding adolescents to the vaccination schedule was found to be crucial in achieving these targets, while the further addition of children ages 5 to 11 was also found to be highly effective in reducing the COVID-19 health burden. Vaccinating children and adolescents has key benefits; it increases the overall population-wide coverage level, and reduces transmission in an age group implicated in high transmission of viruses by aerosol droplets, *e*.*g*. influenza and coronaviruses, in school settings.^4^ Allowing Delta outbreaks to be successfully managed via a combination of achievable coverage rates and moderate physical distancing measures, those that allow schools and many workplaces to remain open, will minimise repeated and enduring hard lockdown measures and resulting economic, health and educational damage. Further research is needed to establish the costs and benefits of such COVID-19 mitigation policies.

In the absence of physical distancing NPI measures, we predict that very high 90% vaccine coverage in those aged 12 and above may result in significant outbreaks of over ∼600,000 cases in a population of 5 million, cities the size of greater Melbourne or greater Sydney, Australia. Furthermore, our modelling suggests that ∼3,400 hospitalisations and almost 500 deaths may result. At the more achievable 80% vaccination coverage, over 850,000 cases may result in the absence of any physical distancing measures, approximately 17% of the modelled population. At its peak we estimate ∼35,000 cases per day, which may result in demand for ∼5000 hospital beds, which is likely to overwhelm healthcare resources.

The addition of moderate NPIs coupled with 80% vaccine coverage in ages 12 and above was found to reduce cases to ∼620,000 in a population of 5 million, ∼13% of the population. This level of vaccination would see peak hospital bed demand at ∼1000 and a total of ∼450 deaths possible. These numbers of cases, hospitalisations and deaths are similar to the reduction which may be achieved at 90% coverage in ages 12 and above. These data suggest that controlling outbreaks of highly transmissible COVID-19 variants without attendant social distancing measures will result in significant numbers becoming infected, unless vaccine coverage significantly greater than 80% can be achieved. The inclusion of children aged 5 to 11 years is shown to reduce the COVID-19 health burden, by increasing the “pool” of immune individuals in the overall population and reducing transmission in school settings. Vaccinating school age children and adolescents has the added advantage that vaccination may be conducted efficiently within schools, as occurs for other infectious diseases in middle/high income countries.

Adoption of moderate NPIs also sees a reductions of 40-65% of cases, hospitalisations and deaths in the elderly for every vaccination coverage level, other than 90% coverage of ages 5 and over, where our data suggests that transmission is effectively eliminated. This older age cohort are at higher risk of poor health outcomes following infection compared to other age groups,^21^ and use of even moderate NPIs should result in a significant reduction in COVID-19 hospitalisations and deaths.

Results provide evidence that moderate physical distancing measures, which importantly permit schools to remain open, can successfully contain future high transmission variants under achievable coverage levels between 80 to 90%. They further highlight the need to activate such NPI measures as early as possible, as found previously.^8^

Results from this study are consistent with a related study which found that vaccine-only strategies were unlikely to achieve herd-immunity in Australia, a study that evaluated vaccination in the context of the less transmissible Alpha variant, with an R_0_ of 2.9.^22^ A further study by the same authors evaluated the current Delta outbreak in Sydney, Australia using a basic reproduction number of 5.97.^23^ That study evaluated the progressive increase in vaccine coverage rates and also suggests the need for concomitant NPI measures to be applied.

The findings are based on a number of assumptions, including the assumed transmission rate of the Delta variant, vaccine effectiveness against transmission, and the timely response of activating physical distancing measures once case numbers are first seen to increase.

Data generated by our model-based analyses may be scaled-up to reflect populations in high-income countries with similar demographics, economies and healthcare systems to Australia, such as New Zealand, those in Europe and North America, and parts of Asia.

## Data Availability

All data referred to is publicly available

## Contributors

GM and DW conceptualised the study. GM and JC designed the model. JC developed the model and generated figures and tables. GM wrote the first draft of the manuscript and verified the data. All authors revised the manuscript critically for important intellectual content, and read and approved the final manuscript version. All authors had full access to all the study data, and the corresponding author had final responsibility for the decision to submit the manuscript.

## Data sharing

All empirical data used in this study are publicly available and have been cited in the Article. The simulation code is available online.

## Acknowledgements

This study was funded by a Future Health Research and Innovation Focus Grant: COVID-19 from the Department of Health, Western Australia.

